# Continuous quality improvement with a two-step strategy effective for large-scale SARS-CoV-2 screening at the Tokyo 2020 Olympic and Paralympic Games

**DOI:** 10.1101/2024.05.29.24307666

**Authors:** Hayato Miyachi, Satomi Asai, Rika Kuroki, Kazuya Omi, Chiaki Ikenoue, Satoshi Shimada

**Affiliations:** Department of Laboratory Medicine, Tokai University School of Medicine, Isehara,259-1193, Japan and Faculty of Clinical Laboratory Sciences, Nitobe Bunka College, Tokyo 164-0001 Japan; Department of Laboratory Medicine, Tokai University School of Medicine, Isehara, 259-1193, Japan; SRL Inc., Tokyo, 107-0052, Japan; SRL Inc., Tokyo, 107-0052, Japan; H.U. Group Research Institute G.K., Tokyo, 197-0833, Japan; and Fujirebio Inc., Tokyo, 107-0052, Japan; Center for Field Epidemic Intelligence, Research and Professional Development, National Institute of Infectious Diseases, Tokyo, 162-8640, Japan; Department of Eco-Epidemiology, Institute of Tropical Medicine (NEKKEN), Nagasaki University, Nagasaki, 852−8523, Japan and Center for Emergency Preparedness and Response, National Institute of Infectious Diseases, Tokyo, 162-8640, Japan

**Keywords:** COVID-19, Olympic Games, sporting events, mass gatherings

## Abstract

The Tokyo 2020 Olympic Games (Games) were held during the height of the coronavirus disease 2019 (COVID-19) pandemic. In order to detect the severe acute respiratory syndrome coronavirus-2 (SARS-CoV-2) early enough to contain the spread as well as to facilitate the timely arrival of athletes at their game venues, all participating athletes up to 14,000 staying in the Olympic Village were screened daily for the infection. Toward this aim, a two-step strategy was adopted comprising screening of self-collected saliva samples using a chemiluminescence enzyme immunoassay followed by confirmatory testing using polymerase chain reaction. The testing system was integrated with an information management system covering all steps. To ensure the accuracy of testing results, rigorous quality assurance measures and monitoring of performance/specimen quality were implemented. The chronological chart analysis was implemented to monitor the holistic process and to give feedback to improve the sampling. Nearly all test results for 418506 saliva samples were reported within 12 hours of sample collection, achieving the target mean turnaround time of 150 minutes for confirmatory testing. As a result, athlete activity and performance for the Games were ensured. The chronological chart confirmed that no athletes were retired due to a false-positive result and no infection clusters among athletes were identified. In conclusion, continuous quality improvement in the two-step strategy for the large-scale screening of COVID-19 contributed to the success of the Games during the pandemic. The quality practice, systems, and workflows described here may offer a model for future mass-gathering sporting events during similar major infectious disease epidemics.

## Introduction

Held in Summer 2021 following a 1-year postponement amid the coronavirus disease 2019 (COVID-19) pandemic, the Tokyo 2020 Games faced the challenge of surging COVID-19 rates, with the incidence reaching unprecedented levels both in Tokyo and globally [1]. The Tokyo 2020 Infectious Diseases Control Centre organized by the Tokyo Organizing Committee of the Olympic and Paralympic Games (TOCOG) played a vital role in containing the spread of severe acute respiratory syndrome coronavirus 2 (SARS-CoV-2) during the Games via the implementation of planned biosafety protocols, including frequent testing for SARS-CoV-2 [2].

In order to detect the virus with sufficient time to contain the spread as well as to facilitate the timely arrival of athletes at their game venues, all participating athletes and others staying in the Olympic Village were screened daily for SARS-CoV-2 infection with a two-step strategy comprising screening of self-collected saliva samples using a chemiluminescence enzyme immunoassay (CLEIA) followed by confirmatory testing of saliva and nasopharyngeal swab samples using polymerase chain reaction (PCR) [2,3] instead of PCR screening. Although the “gold standard” of SARS-CoV-2 testing via PCR using NP swab samples is accurate and reliable, results may take 24–48 h to return. Such delays may lead to not only further transmission of disease but can also diminish the daily activity of athletes for training and arrival at the match venue. Furthermore, it requires trained professionals in full protective equipment to collect specimens one person at a time. Solutions to improve the efficiency of mass-screening for SARS-CoV-2 include the replacement of NP swab samples with self-collected saliva thereby eliminating specialized medical personnel and allowing simultaneous parallel sample collection [3]. To monitor in real-time the workflow processes of each individual test, the testing system was integrated with an information management system covering all steps from specimen collection to reporting results. This strategic allocation of PCR tests to cases that were CLEIA-positive or inconclusive not only maintained accuracy but also significantly improved logistical efficiency in administering mass screenings. Notably, PCR testing is associated with a potential risk of false-positive results, which causes unnecessary withdrawal of athletes from the Games. In order to ensure the COVID-19 screening test, we implemented continuous quality improvement through rigorous quality assurance measures, performance/specimen quality monitoring by an internal quality assessment, and a chronological chart analysis for individual cases.

## Material and methods

### Patient and public involvement

The study’s design including participants was planned and conducted under requests and the endorsement of the TOCOG according to the regulation and the criteria specified by the Ministry of Health, Labour, and Welfare of Japan. Up to 14,000 athletes and team officials were tested daily for SARS-CoV-2 in order to detect infections early as required by the TOCOG.

### A two-step large-scale screening strategy

To facilitate the timely arrival of athletes at their game venues, test results had to be available within a 12-hour turnaround time (TAT). For this large-scale screening, a two-step strategy was adopted, instead of PCR screening [2,3]. First, quantitative antigen testing was performed using a quantitative CLEIA with a fully automated instrument (Lumipulse® Presto SARS-CoV-2 Ag, Fujirebio, Tokyo, Japan). The quantitative CLEIA is a high-throughput assay with high sensitivity, specificity, and short TAT, and can provide rapid results as an initial saliva screening test. Positive or inconclusive results were confirmed using a standard PCR test. If the saliva PCR showed positive results, a nasopharyngeal (NP) swab was collected at the athletes’ village fever clinic for confirmatory PCR testing using the SARS-CoV-2 Direct Detection RT-qPCR Kit (Takara Bio Inc., Kusatsu, Japan) with instruments such as the QuantStudio5 (Thermo Fisher Scientific K.K., Waltham, MA, USA), LightCycler 96 (Roche Molecular Systems, Inc., Pleasanton, CA), and CFX96 (Bio-Rad Laboratories, Inc., Hercules, CA, USA). Symptomatic participants also underwent PCR testing of NP swabs. The workflow was integrated with an information management system. Self-collected saliva was brought to the reception area by COVID-19 Liaison Officers (CLOs). At the reception department, accreditation ID was linked with the sample ID in the information management system by barcode. Radiofrequency identification (RFID, Toppan Forms Inc., Tokyo, Japan) was incorporated into the information management system for specimens. The laboratory information and reporting system (Neo Polaris and Fukuda System, H.U. Group Research Institute G.K., Tokyo, Japan) allowed for real-time monitoring of the workflow processes of each test, covering the entire process from receipt of the specimen to reporting the result to CLOs in addition to recording the internal laboratory workflow (Figure 1).

**Fig 1.**
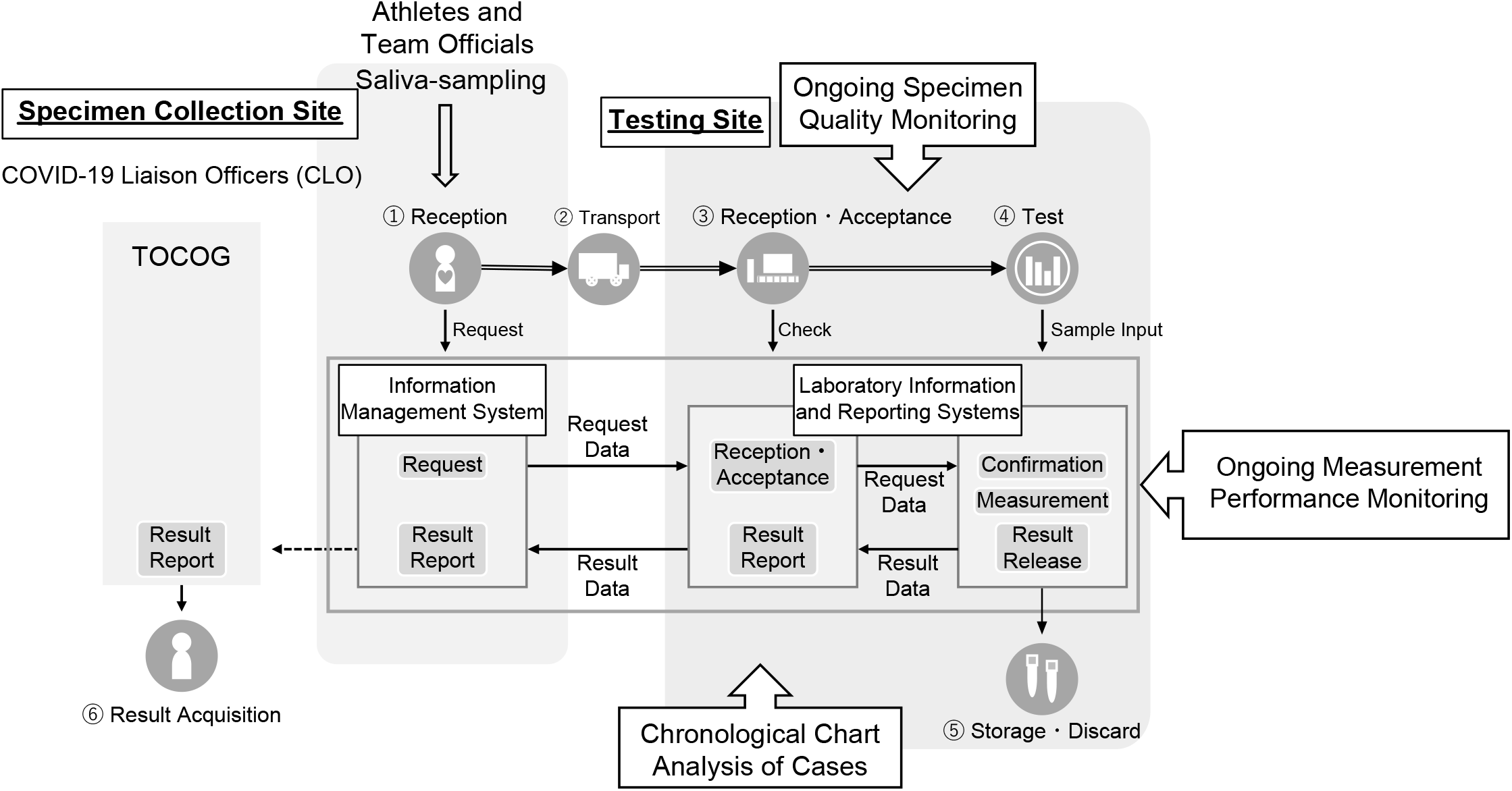
Workflow Quality Monitoring. The workflow was integrated with an information management system. Self-collected saliva was brought to the reception area by COVID-19 Liaison Officers (CLOs). The accreditation ID of personnel for the Village was linked with radiofrequency identification and sample ID barcodes. This feedback process was used to communicate with the Tokyo Organizing Committee of the Olympic and Paralympic Games (TOCOG) about re-training of CLOs and TOCOG staff in appropriate collection techniques, including self-sampling of saliva before eating, specimen refrigeration within 24 hours of sampling, and NP swab collection, enabling real-time corrections of the process.

Five laboratories coordinated the conducting of testing for various Olympic Games venues. Quantitative CLEIA and PCR tests were also performed in four laboratories outside the village, located in Kawasaki, Tokyo, Sapporo, and Iwaki.

### Rigorous quality assurance measures

As the Tokyo 2020 Games were an international multi-sport event during a pandemic, quality laboratory practices based on both global and regional perspectives were rigorously implemented [4,5]. Actions taken were based on discussion and consensus during the development of the International Organization for Standardization standard (ISO/TS 5798) for SARS-CoV-2 detection using nucleic acid amplification methods [4]. To maintain the expected performance of SARS-CoV-2 laboratory tests, meticulous quality assurance measures such as personnel training, competency assessments, assay performance evaluations, and internal quality controls based on cycle threshold values, were implemented. The assay performance was monitored and confirmed by a nationwide external quality assessment of SARS-CoV-2 nucleic acid amplification tests in Japan [5].

## Results

### Test volume and turnaround time

During the Games, all participating athletes up to 14,000 and others staying in the Olympic Village were screened daily for SARS-CoV-2 infection with a two-step strategy comprising screening of self-collected saliva samples using a CLEIA followed by confirmatory testing of saliva and nasopharyngeal swab samples using PCR. Using a testing system integrated with an information management system covering all steps, from specimen collection to reporting of the results, enabled monitoring processes of each testing down to confirmatory test results to be provided. A total of 418,506 saliva samples were tested using the quantitative antigen test. Nearly all tests were reported within 12 hours of sample collection, aligning with the target of a mean TAT of 150 minutes for confirmatory tests.

### Continuous quality improvement

Quality assurance of tests based on both global and regional perspectives was rigorously implemented. To ensure the entire examination process from sample collection to the results report, we developed and analyzed a chronological chart of cases with positive and inconsistent results, providing a holistic overview of the testing process, confirming test results, and identifying underlying problems including poor specimen quality and false-positive results. Inconsistent results were interpreted and judged whether reasonable as a natural course of infection or suspicious test result due to a possible error in any portion of the testing process.

Particularly, sample collection methods could impact the results; the pre-examination process posed a critical challenge for mass screening. CLOs played a crucial role in instructing athletes and team officials in proper self-sampling procedures. However, self-sampling in individual rooms had a risk of poor specimen quality due to suboptimal collection and storage practices. Visual examination of specimens revealed sediments, which caused nonspecific reactions, increasing rates of re-measurement, and prolonged the TAT. To mitigate these nonspecific reactions due to sediments, the phosphate-buffered saline for dilution was changed to a proprietary buffer. We evaluated specimen quality using amylase measurement with CicaLiquid AMY (Kanto Chemical Co., Inc., Tokyo, Japan) on a fully automated analyzer (JCA-BM8040, JEOL Ltd., Tokyo, Japan). A high proportion of specimens showed amylase levels under the reference range, suggesting inappropriate storage of specimens at ambient temperature.

In the continuous quality improvement of testing during the Games, continuous monitoring was implemented to identify potential bias, which allowed real-time actionable feedback to the TOCOG. We used this feedback process to communicate with TOCOG regarding the re-training of CLOs and TOCOG staff in appropriate collection techniques including self-sampling of saliva before eating, specimen refrigeration within 24 hours of sampling, and NP swab collection. This enabled real-time corrections in the testing process. As a result, the activity and performance of athletes for daily training and participation in the games were ensured. The chronological chart for the individual test results enabled us to confirm in real-time that there were neither infection clusters identified in athletic teams inside the village or Games venues due to false-negative results, nor unnecessary withdrawal of athletes from the Games due to false-positive results.

### Limitations

The testing for SARS-CoV-2 detection to detect infections early was performed, according to the regulation and the criteria specified by the Ministry of Health, and Labour in Japan. Ideally, the quality and competence of the laboratory and the reliability of its testing service would be objectively audited and endorsed by the third-party accreditation. However, in Japan, laboratory accreditation under ISO 15189, Medical Laboratories – Requirements for quality and competence [6], applies laboratory tests for medical use but not those for public use such as for SARS-CoV-2 screening tests of participating athletes and others staying in the Olympic village. Thus, the laboratory was not subjected to accreditation under ISO 15189. The operation of laboratories was built and conducted, in alignment with the criteria specified for the central laboratory which had been accredited under ISO 15189 and had a responsibility for the testing for SARS-CoV-2 detection under a contract with TOCOG.

## Discussion

In order to detect SARS-CoV-2 infections as early as possible and to take measures to prevent further spread, a two-step strategy was adopted to meet the large-scale screening requirements of the event. The integration of a testing system with an information management system covering all steps from specimen collection to reporting of the results enabled confirmatory test results to be provided within 12 hours, achieving our target of a mean TAT of 150 minutes for confirmatory tests. This integration enabled all athletes to keep the daily activity of training prior to the match and participate at the game match venue.

The accuracy of testing results can have an impact on the risk of transmission of disease. False-negative results may lead to further transmission of disease. False-positive results would lead to the withdrawal of the athlete from further participation in the games. Considering the Tokyo 2020 Games being an international multi-sport event during a pandemic, quality laboratory practices based on both global and regional perspectives were rigorously implemented. In order to minimize such a risk, we implemented and maintained rigorous quality assurance of the entire process of examination of daily testing for SARS-CoV-2. Quality assurance measures included personnel training, competency assessments, assay performance evaluations, and internal quality controls based on cycle threshold values. In addition to that, the pre-examination process was a critical challenge of mass screening in the event. CLOs instructed athletes and team officials on how to perform self-sampling. However, self-sampling in individual rooms posed a risk of poor specimen quality due to suboptimal collection and storage. In order to monitor the quality of specimens, we adopted visual inspection and measurement of amylase in saliva.

This shows that international major sporting events could be held during major infectious disease epidemics comparable to the COVID-19 pandemic without causing spikes in the case numbers [7]. These results support the approach, advocated by the World Health Organization, that responding to and managing the COVID-19 pandemic requires using all available options, including public health and social measures, robust test-and-trace systems, and vaccination [1]. During the early phase of the COVID-19 pandemic, most mass-gathering sports events used PCR testing for infection control, which required an extended TAT for receiving results and enforcing quarantine [8-10]. Furthermore, PCR has a potential risk of false-positive results, which caused unnecessary withdrawal of athletes from the Games. Our experience suggests that continuous quality improvement implemented in the two-step strategy and infection control measures at the Games were successful.

The SARS-CoV-2 positivity rate among the 11,417 Olympic and 4,393 Paralympic athletes was 0.03% of the total tests conducted [11]. Notably, the chronological chart for the individual test results enabled us to confirm in real-time that there were neither infection clusters identified in athletic teams inside the village or Games venues due to false-negative results, nor unnecessary withdrawal of athletes from the Games due to false-positive results.

## Conclusion

In conclusion, continuous quality improvement was implemented in the two-step strategy for large-scale COVID-19 screening and contributed to the success of international major sporting events during the pandemic. The quality practice, systems, and workflows of testing with continuous quality improvement, described in this report could serve as a model for future mass-gathering sporting events during similar major infectious disease epidemics.

## Data Availability

The data that support the findings of this study are available from the corresponding author upon reasonable request.

## Author Contributions

Conceptualization and design, H.M., S.A., R.K., K.O., C.I., and S.S.; material preparation, data collection and analysis, H.M., R.K., and K.O., and S.S.; writing— review and editing, H.M., S.A., C.I., and S.S. All authors commented on previous versions of the manuscript. All authors read and approved the final manuscript.

### Institutional Review Board Statement

Ethical review and approval were waived for this study due to the Act on the Prevention of Infectious Diseases and Medical Care for Patients with Infectious Diseases (the Infectious Diseases Control Law) in Japan.

### Informed Consent Statement

Patient consent was waived due to the Act on the Prevention of Infectious Diseases and Medical Care for Patients with Infectious Diseases (the Infectious Diseases Control Law) in Japan.

## Acknowledgments

We would like to thank the Tokyo 2020 Organizing Committee. The findings and conclusions in this report are those of the authors and do not necessarily represent the official position of the Tokyo 2020 Organizing Committee. The Tokyo 2020 Organizing Committee does not endorse any commercial products or services.

